# Differences in tuberculosis prevalence among people living with and without HIV in low-and-middle-income countries: A systematic review and meta-analysis

**DOI:** 10.64898/2026.04.20.26351343

**Authors:** Nicole A. Swartwood, Melike Hazal Can, Seyed Alireza Mortazavi, Hening Cui, Nanki Singh, Do Kyung Ryuk, Katherine C. Horton, Peter MacPherson, Nicolas A. Menzies

**Affiliations:** Department of Global Health and Population, Harvard TH Chan School of Public Health, Boston MA, USA; School of Health and Wellbeing, University of Glasgow, Glasgow, United Kingdom; Department of Population Health, NYU Grossman School of Medicine, New York, NY, USA; Division of Research, Kaiser Permanente Northern California, Pleasanton CA, USA; Department of Infectious Disease Epidemiology, London School of Hygiene and Tropical Medicine, London, United Kingdom; Center for Health Decision Science, Harvard T H Chan School of Public Health, Boston, MA, USA

## Abstract

**Background:** Tuberculosis (TB) and human immunodeficiency virus (HIV) are leading causes of infectious disease deaths, with disproportionate impact in low- and middle-income countries (LMICs). Despite well-established biological relationships between these diseases, there is limited information on how TB prevalence differs between people living with and without HIV.

**Methods:** We conducted a systematic review and meta-analysis of TB prevalence surveys conducted in LMICs and published during January 1^st^ 1993–October 13^th^ 2025 (PROSPERO CRD42024503853). We extracted bacteriologically-confirmed TB prevalence estimates stratified by participant HIV status. Surveys that offered HIV testing to all, sputum-collection-eligible, or TB-positive participants were included in the primary analysis. We applied Bayesian meta-regression to estimate pooled risk ratios (RR) of bacteriologically-confirmed TB prevalence among participants living with versus without HIV. Additionally, we estimated country-level and overall TB notification-to-prevalence (N:P) ratios by HIV status.

**Findings:** Of 10,211 potentially relevant publications, 12 TB prevalence surveys—representing 264,530 participants within nine countries in Southern and Eastern Africa—were used in the primary analysis. Reported TB prevalence was higher among participants living with versus without HIV in 11/12 surveys, with an overall pooled RR of 3·86 (95% credible interval: 2·41–5·53). N:P ratios were higher among participants living with HIV in all examined countries. The overall pooled N:P ratios were 1·74 (0·59–4·56) and 0·48 (0·17–1·20) among participants living with versus without HIV, respectively.

**Interpretation:** In Southern and Eastern Africa, bacteriologically-confirmed TB prevalence is three- to six-times higher among people living with HIV. Comparison of prevalence and notification data suggest higher rates of TB diagnosis for people living with versus without HIV, but also indicates substantial delays in the detection of untreated TB cases for both populations.

**Funding:** Wellcome Trust, UK National Institute for Health and Care Research, UK Foreign, Commonwealth and Development Office, NIH.

**Research in context:** *Evidence before this study:* There is limited systematic evidence on how the prevalence of TB disease differs between people living with HIV and without HIV. Multiple observational cohorts have described substantially elevated TB incidence among populations with HIV, but disease prevalence will also be affected by differences in mortality and treatment uptake rates. We searched PubMed from inception through January 21, 2026 using the search string ((HIV AND TB) OR HIV/TB) AND (prevalence AND (systematic review OR meta-analysis)) without any restrictions on language. We also reviewed investigators’ personal libraries. This search yielded 506 publications; however few of these included prevalence data. An analysis conducted in 2020 synthesized HIV status-stratified data from seven national TB prevalence surveys in Africa and found that HIV prevalence was lower among prevalent TB cases than among notified cases. This study did not include subnational surveys and did not distinguish between survey participants with self-reported or test-confirmed HIV status.

*Added value of this study:* This study synthesized TB prevalence data, stratified by participant HIV status, from national and subnational surveys conducted in LMICs and published between January 1^st^ 1993 and October 13^th^, 2025. Collated data represented 681,402 survey participants across ten countries. All but one study were conducted in Southern and Eastern Africa. We limited our primary analysis to surveys that systematically tested participants for HIV and bacteriologically-confirmed TB. The prevalence of bacteriologically-confirmed TB was estimated to be three to six times higher than among people living with versus without HIV. Ratios of TB notifications to TB prevalence were higher for people living with HIV compared to people without HIV, suggesting higher rates of TB case detection (and likely shorter duration of disease) for people living with HIV and untreated TB than those without HIV.

*Implications of all available evidence:* Few estimates of community-representative TB prevalence stratified by participant HIV status exist. These surveys have been concentrated in Southern and Eastern Africa, despite TB-HIV burden being distributed globally. Our findings highlight the elevated burden of TB among people living with HIV in these settings, as well as the limited data on the intersection of TB and HIV epidemiology in other world regions. Furthermore, our comparison of notification and prevalence data demonstrate substantial shortfalls in TB case detection, regardless of an individuals’ HIV status.

## Introduction

Tuberculosis (TB) and human immunodeficiency virus (HIV) are leading causes of infectious disease deaths, disproportionately impacting low- and middle-income countries (LMICs).^1^ HIV affects TB epidemiology in multiple ways, with people living with HIV more likely to develop TB disease following *Mycobacterium tuberculosis* infection,^2^ and those that develop TB facing more rapid disease progression, with higher risks of disseminated and extra-pulmonary TB.^3^ People living with HIV also experience higher TB-related mortality than people living without HIV,^4^ with 15% of global TB deaths in 2024 occurring among HIV co-infected individuals.^1^ These elevated TB risks are mediated by immune function, increasing with progressive HIV-associated immunosuppression, and decreasing following the receipt of antiretroviral therapy (ART).^5^

Given the intertwined epidemiology of these diseases, it is valuable to understand the relative burden of TB among people living with versus without HIV. However, differences in TB burden by HIV status cannot be accurately estimated directly from routinely reported data. A number of factors affecting TB case ascertainment—disease presentation,^3^ access to and frequency of clinical care,^6,7^ performance of diagnostic tests,^8^ and clinician suspicion of TB^9^—differ systematically between people living with versus without HIV, such that HIV prevalence among individuals diagnosed with TB (as captured by TB case notifications) provides a distorted picture of the differential risk of TB by HIV status.

Given the biases inherent in TB case notifications, TB prevalence surveys are seen as a critical source of evidence for quantifying the underlying disease burden in communities. As these surveys attempt complete population screening, they may be unaffected by the biases associated with notification data, such as underlying variation in healthcare access, case detection, and incomplete reporting. If sample sizes are large enough, community-based survey data can be disaggregated by key population characteristics to elucidate differential TB burden across the sampled population, which may increase the resources needed to implement the survey. For example, to describe differences between people living with and without HIV, surveys must provide HIV testing for study participants or, less reliably, assess participant HIV status via self-report.

As TB prevalence surveys are resource intensive and conducted infrequently, population-representative prevalence estimates might not be available for all settings of interest. Consequently, there is value in synthesizing available survey data to understand systematic trends and patterns that apply across settings. To better understand the differences in TB burden by HIV status, we conducted a systematic review and meta-analysis of TB prevalence surveys that were conducted in LMICs and reported TB prevalence estimates disaggregated by participant HIV status. Additionally, we compared these prevalence data to similarly disaggregated TB notifications data, to draw inferences about relative rates of TB diagnosis among people with untreated TB. The resulting notification-to-prevalence ratios can provide a useful tool for understanding the rate at which individuals with prevalent TB receive a diagnosis, and can be used to identify gaps in TB detection and reporting.^10^

## Methods

We conducted a systematic review and meta-analysis to estimate TB prevalence stratified by participant HIV status in LMICs (as defined by 2024 World Bank classification).^11^ This review was prospectively registered with the International Prospective Register of Systematic Reviews (PROSPERO, protocol number CRD42024503853). We conducted this analysis in accordance with Meta-analysis of Observational Studies in Epidemiology (MOOSE) guidelines and prepared the manuscript to align with the Preferred Reporting Items for Systematic Reviews and Meta-analyses (PRISMA) guidelines.^12,13^ The review was managed with the Covidence online systematic review tool.^14^

### Search strategy

We searched four electronic databases—the Cochrane Database of Systematic Reviews, Embase, PubMed, and Global Health—for English-language publications with a publication date between December 1, 2015 and October 13, 2025 (full search strategy reported in Table S1). We also reviewed prevalence surveys included in a previous systematic review, published in the WHO Global Tuberculosis Report 2025, or described in abstracts from the 2023 Union World Conference on Lung Health.^1,15,16^ Finally, we examined the citations of all identified publications for potentially relevant publications.

### Selection criteria

Our review team was comprised of nine people (NAS, MHC, AM, HC, NS, DKR, PM, KCH, NAM). Two reviewers (of NAS, MHC, AM, HC, DKR, assigned at random) independently reviewed titles and abstracts of all studies to identify those meeting the criteria for full text review. Once identified, two reviewers (of NAS, MHC, AM, HC, DKR, KCH, PM) reviewed each full text for study inclusion. In each of these steps, discrepancies were resolved through discussion between the initial reviewers, mediated by a third reviewer (NAS, PM, KCH, or NAM).

We included surveys that reported adult (≥15 years) pulmonary TB prevalence from a population-based sample, disaggregated by participant HIV status. We excluded surveys that included only pediatric (age <15 years) populations, extra-pulmonary TB, or drug-resistant TB, due to the difficulty of community-based detection of these conditions. We also excluded studies that only included individuals seeking care, assessed TB status via self-report, or reported the results of surveys conducted after an active case finding intervention in the same setting. Table S2 reports detailed inclusion and exclusion criteria.

Two members of the study team independently extracted data on each survey’s sampling methodology, HIV and TB diagnostic algorithms, risk of bias (described below), and HIV-stratified TB prevalence using a pretested extraction form (Form S1). Extraction discrepancies were resolved by a third reviewer.

### Risk of bias

We assessed each study’s risk of bias through eight criteria adapted from a tool developed by Hoy et al (2012) for the appraisal of prevalence surveys.^17^ These criteria evaluated study population selection, data collection methodology, participant non-response bias, and case definition parameters. Full bias assessment criteria are included in the extraction form (Form S1, Section 8). Each reviewer also provided a summary assessment of each study’s overall risk of bias (low, moderate, or high) based on the evaluated criteria.

To explore potential language bias, we replicated our search and review process for publications published in Spanish, Portuguese, or French. We searched three electronic databases—LILACS, SciELO, and Africa Index Medicus. Table S4 presents the detailed search strings. We used deepL Translate, a neural machine translation service, to translate the titles, abstracts, and identified full texts into English for review.

### Definitions

A survey participant was defined as a person who provided consent for inclusion and underwent initial TB screening (usually symptom screening and/or chest radiography). We defined bacteriologically-confirmed TB as TB disease that was diagnosed following a positive result on sputum smear microscopy, culture, and/or a molecular diagnostic such as Xpert MTB/RIF. We used each study’s definition of bacteriologically-confirmed TB if reported. Otherwise, we used estimates of exclusively culture-positive or smear-positive TB prevalence, prioritized in that order. Smear-positive TB was defined as TB confirmed through sputum smear microscopy, irrespective of other diagnostic results. If a study reported more than one TB prevalence measure, adjusted TB prevalence estimates with 95% confidence intervals were the preferred measure used for synthesis, followed by crude prevalence estimates with 95% confidence intervals, then counts of individuals with TB and total number tested, or summary prevalence estimates without a confidence interval (Figure S1).

### Data analysis

To be considered for primary quantitative analysis, we required the survey to include a systematic offer of HIV testing to participants, i.e. testing offered to all participants, all participants eligible for sputum collection, all TB positive participants, etc. Surveys that ascertained HIV status solely through participant self-report were analyzed as a secondary outcome. If a survey offered HIV testing but recorded self-reported HIV status if testing was refused, it was included in primary analysis if <10% of known HIV status was ascertained via self-report. Finally, if a survey excluded participants on ART from HIV testing, but provided testing to all remaining participants, it was included in primary analysis, with participants receiving ART classified as HIV-positive.

For surveys that only offered HIV testing to either participants with presumptive (sputum-collection eligible) or diagnosed TB, we estimated HIV prevalence among the screened population by multiplying total participant counts by external HIV prevalence estimates matched to survey geography and end year.^18^ We used these denominator estimates and study reported data on HIV-status-stratified TB cases to estimate TB prevalence among people living with and without HIV.

### Risk ratio of TB prevalence among people living with and without HIV

Our primary outcome was the risk ratio of bacteriologically-confirmed TB prevalence among people living with HIV compared to those without HIV, estimated using Bayesian multi-level regression models. Model equations and parameter assumptions are reported in Table S4. In the primary analysis, we fit a regression model with study- and country-level nested random effects to data on bacteriologically-confirmed TB from surveys with test-confirmed HIV status.

To examine how testing algorithms affect the prevalence risk ratio, we re-fit the regression model to data representing (a) smear-positive TB prevalence among surveys that evaluated HIV status via survey-provided testing and (b) bacteriologically-confirmed TB prevalence among surveys that evaluated HIV status via self-reporting. To evaluate the potential relationship between ART coverage and the risk ratio of bacteriologically-confirmed TB prevalence, we extended the regression model for the primary outcome to include a covariate for country-level ART coverage, matched to survey country and year.^18^

### Rate ratio of TB notifications among people living with and without HIV

To calculate HIV-status-stratified notification rates, we collated country- and year-specific data on reported total TB notifications stratified by HIV-status,^19^ as well as total and HIV-positive population estimates.^18^ We included only country-year pairs corresponding to a nationally-representative prevalence survey (N=5) in our primary analysis. Using these data we fit a model with country-level random effects to estimate the pooled rate ratio of TB notifications (TB notification rate for people living with HIV divided by notification rate for people living without HIV).

### Notification-to-prevalence ratios among people living with and without HIV

We fit a multivariate regression model with country-level random effects to jointly estimate the country-level risk ratio of TB prevalence and rate ratio of TB notifications as correlated outcomes for people living with and without HIV. These models were fit to data from the five nationally-representative surveys and country- and year-matched notifications. We computed country-specific notification-to-prevalence (N:P) ratios stratified by HIV status from the two modelled outputs. We also extended this model to include country-level ART coverage, allowing for differential effects by HIV status.

### Statistical analysis

All analyses were performed in R (version 4.4.2) using the ‘brms’ and ‘tidybayes’ packages.^20–22^ Models ran for 10,000 iterations (2,500 burn-in) on four chains. Convergence was assessed through inspection of effective sample size (ESS), potential scale reduction factor (R^), and posterior predictive checks. For all outcomes, we report the posterior mean and equal-tailed 95% credible interval obtained from summarizing 30,000 posterior draws per parameter. The extracted dataset, clean dataset, and analysis code are available at https://github.com/nswartwo/tb-lmic-prevalence-review/ and archived on Zenodo.

## Results

Our search strategy identified 10,124 unique publications that were screened by title and abstract. Among these, 201 publications had their full text reviewed for HIV-stratified TB prevalence estimates (Figure 1). From this review, we identified 17 unique community-representative TB prevalence surveys that reported at least one TB prevalence measure stratified by HIV status.^23–54^ Publications excluded at the full text review stage are listed in Table S5, with their exclusion reason. The primary exclusion reason was the lack of TB prevalence estimates stratified by HIV status (N=98; 49%).

**Figure 1:**
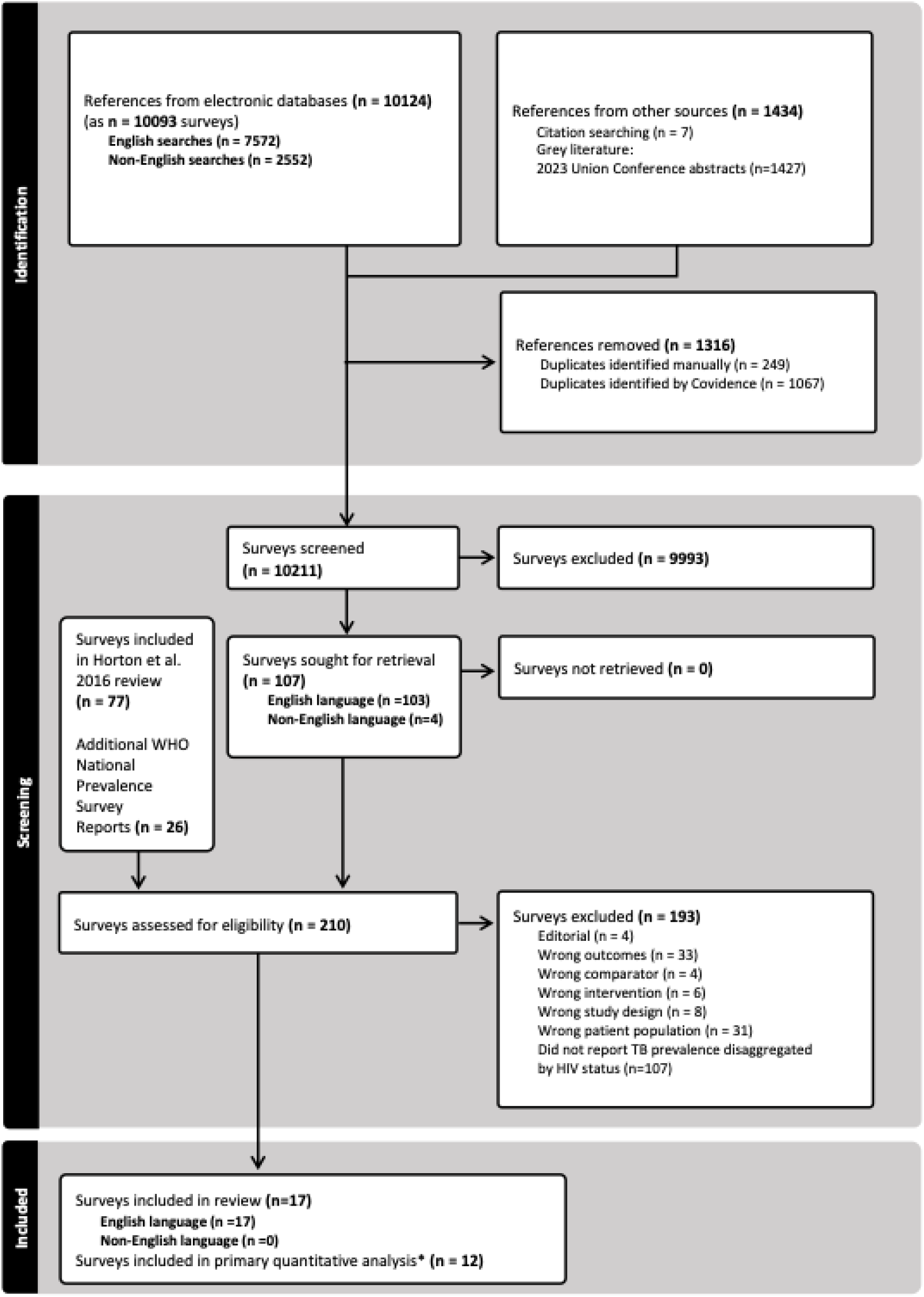
PRISMA diagram of study selection.

All included studies were English-language publications. The extracted surveys represented ten countries: nine in the WHO Africa region and one in the South East Asia region (Figure 2A). Survey data collection ranged from 2005–2022, with nationally-representative surveys conducted during 2012–2019 (Figure 2B).

**Figure 2:**
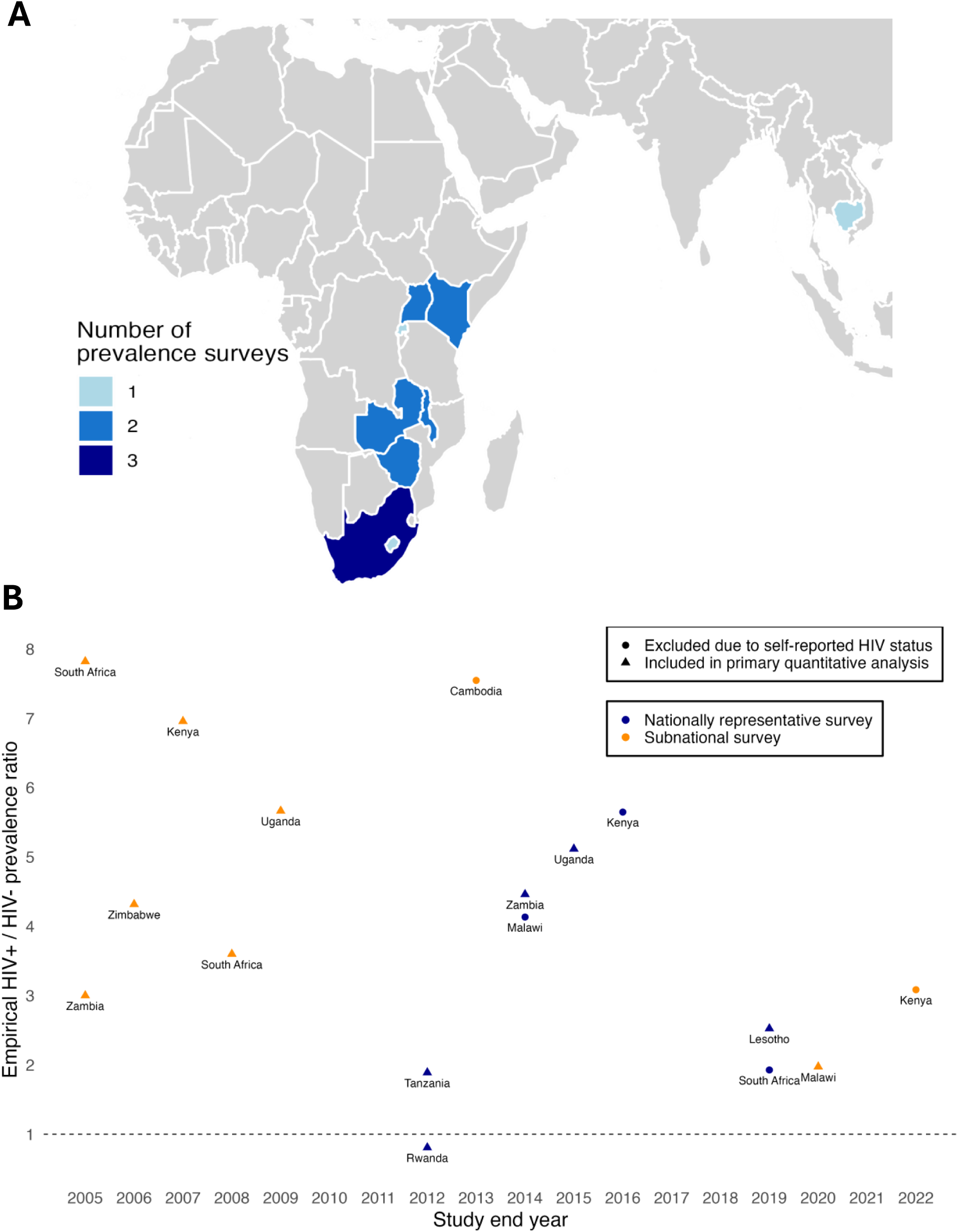
Geographic and temporal distribution of TB prevalence surveys reporting HIV-status stratified estimates. Panel A: Geographic distribution of estimates. Panel B: Distribution of surveys by data collection end year.

**Figure 3:**
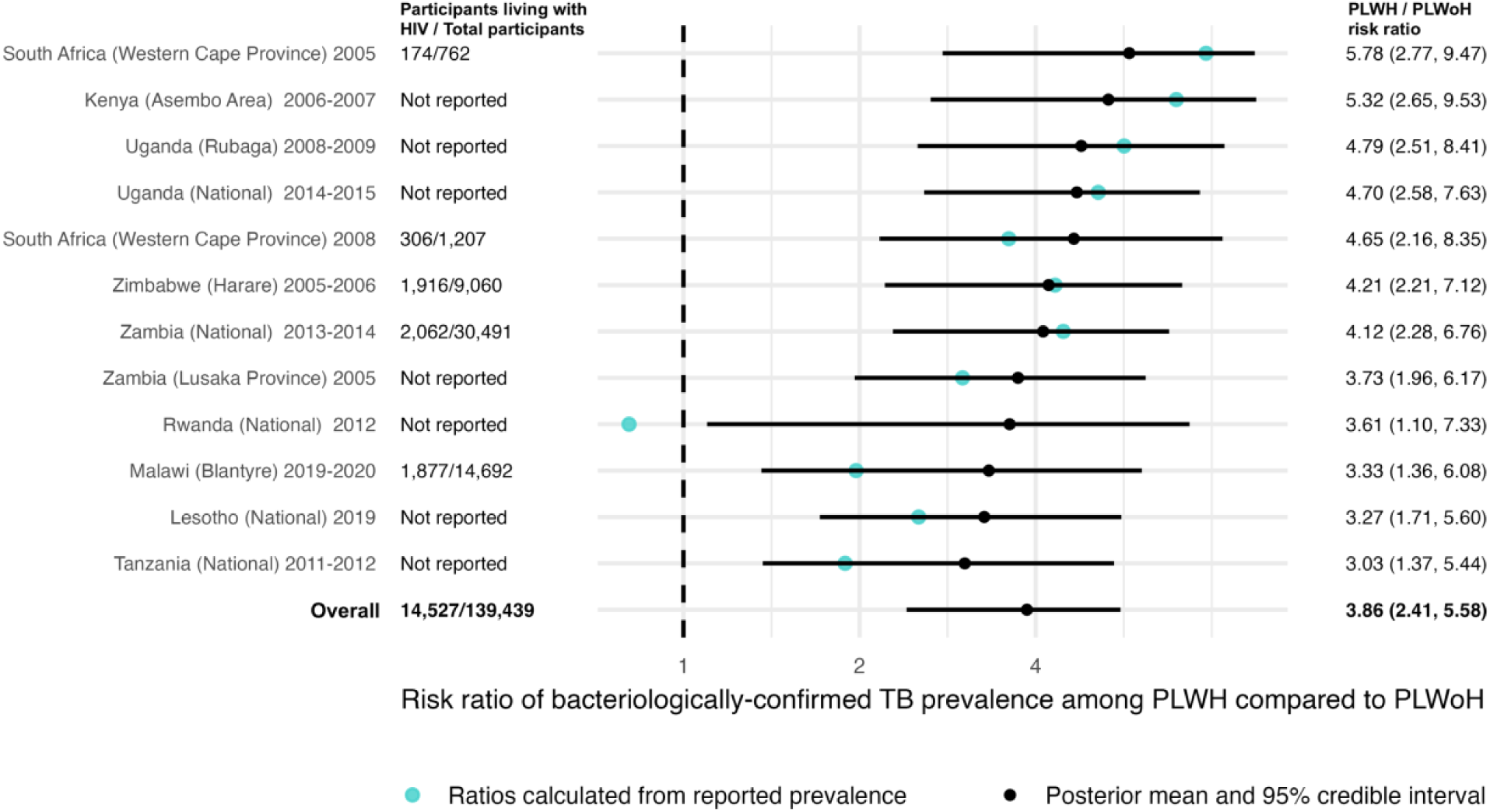
Risk ratio of bacteriologically confirmed TB prevalence among people with and without HIV for each quantitively analyzed survey and overall pooled estimate. Note: Values in parentheses represent 95% credible intervals. PLWH: people living with HIV. PLWoH: people living without HIV.

We included 12 surveys in the primary quantitative analysis, representing 9 countries. Of these, four had denominators estimated from external HIV prevalence data (Table 1). The included surveys varied in their approach to HIV testing. For one survey—Lesotho 2019—HIV status was largely ascertained by HIV testing, but for a small percentage of participants (5%; 884/17,915) was elicited through self-report.^26,43^ No survey reported differential HIV testing procedures for people on ART. In total, the 12 surveys included 264,530 participants. HIV status was known for 97,357 participants (37%), of whom 11,398 (12%) were HIV-positive. Comparatively, the five surveys which collected HIV status via participant self-report included 416,872 participants, three of which were nationally representative. Summary characteristics of survey participants across the TB screening and diagnostic cascade are presented in Table S6.

**Table 1:**
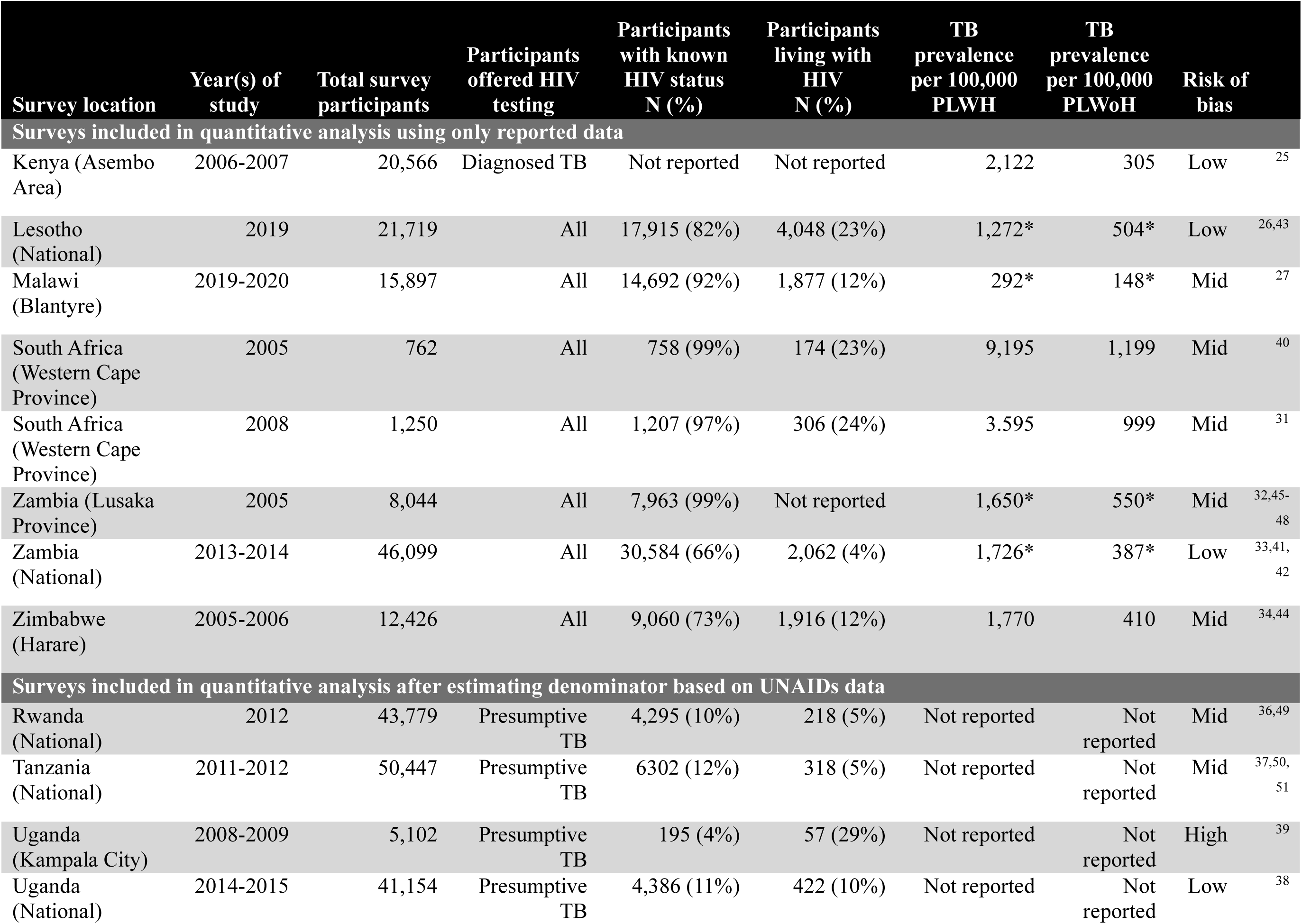
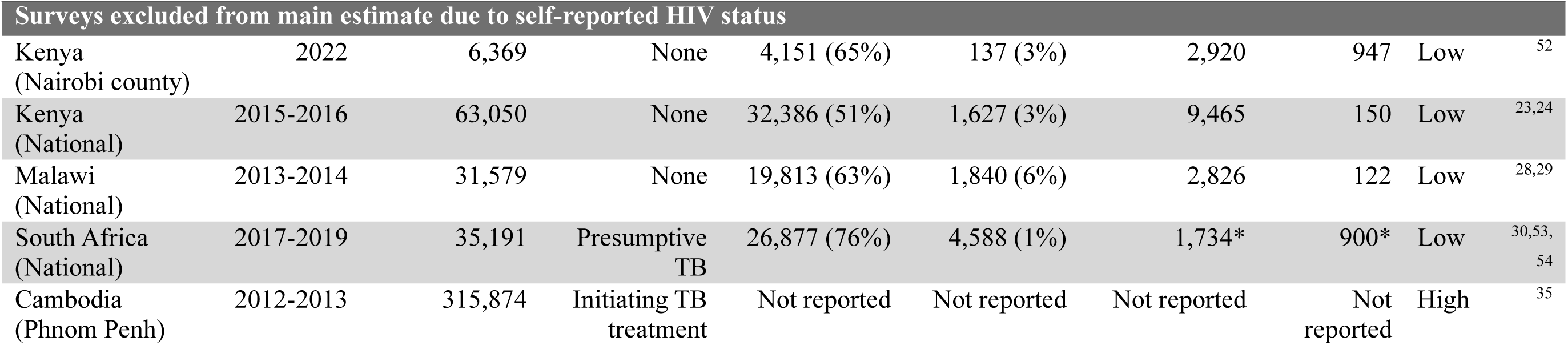
Identified surveys which reported HIV-status stratified TB prevalence estimates. Notes: Prevalence estimates marked with * are study-reported adjusted TB prevalence estimates. Studies with no reported prevalence estimates reported counts of TB cases stratified by HIV status but did not report the associated HIV status stratified participant numbers. For these studies, estimates of country-level HIV prevalence were sourced from UNAIDS and used to calculate HIV population estimates for calculation of TB prevalence rates. PLWH: people living with HIV. PLWoH: people living without HIV.

**Table 2:** Estimated differences in nationally-representative TB prevalence and TB notifications among people living with and without HIV for each country included in main analysis. Note: Values in parentheses represent 95% credible intervals. Analyses marked with (*) included only countries with prevalence estimates provided by nationally-representative surveys. PLWH: people living with HIV. PLWoH: people living without HIV.

| **Survey country** | **PLWH/**<br>**PLWoH**<br>**prevalence**<br>**ratio** | **PLWH/**<br>**PLWoH**<br>**notification**<br>**ratio\*** | **PLWH**<br>**notification-to-**<br>**prevalence**<br>**ratio\*** | **PLWoH**<br>**notification-to-**<br>**prevalence**<br>**ratio\*** |
| --- | --- | --- | --- | --- |
| **Lesotho** | 3.27<br>(1.71, 5.60) | 9.86<br>(3.65, 19.90) | 1.51<br>(0.57, 3.39) | 0.43<br>(0.16, 0.96) |
| **Rwanda** | 3.61<br>(1.10, 7.33) | 12.20<br>(4.19, 23.10) | 2.37<br>(0.53, 6.71) | 0.66<br>(0.16, 1.76) |
| **Tanzania** | 3.03<br>(1.37, 5.44) | 12.20<br>(4.13, 23.00) | 1.74<br>(0.68, 3.82) | 0.49<br>(0.20, 1.04) |
| **Uganda** | 4.70<br>(2.58, 7.63) | 11.30<br>(4.04, 21.50) | 1.48<br>(0.59, 3.13) | 0.42<br>(0.17, 0.88) |
| **Zambia** | 4.12<br>(2.28, 6.76) | 12.70<br>(4.27, 24.50) | 1.42<br>(0.55, 3.18) | 0.41<br>(0.16, 0.88) |
| **Overall** | 3.86<br>(2.41, 5.58) | 11.60<br>(4.01, 22.60) | 1.70<br>(0.58, 4.42) | 0.48<br>(0.17, 1.19) |

Four surveys—Kenya 2006–2007,^25^ Malawi 2019–2020,^27^ Zambia 2013–2014,^33,41,42^ and Zimbabwe 2005–2006^34,44^—reported smear-positive TB prevalence stratified by HIV status (in addition to a separate definition of bacteriologically-confirmed TB). These surveys had a combined 92,798 participants, of whom 74,809 had known HIV status and 9·4% were HIV-positive.

Based on the 12 surveys included in the primary analysis, the pooled estimate of bacteriologically-confirmed TB prevalence was four-times higher in people living with versus without HIV (risk ratio [RR] 3·86; 95% credible interval 2·41–5·53). In the five surveys that collected self-reported HIV status, the pooled RR of bacteriologically-confirmed TB prevalence was 4·00 (1·28–8·19) among people living with vs. without HIV. In the four surveys that reported smear-positive TB prevalence, the pooled RR of smear-positive TB prevalence was 4·21 (0·13–19·53), compared to the pooled RR of 4·02 (0·54–11·45) for bacteriologically-confirmed TB estimated from the same four surveys.

Covariate analysis estimated a 25·1% (−30·7%–61·4%; no statistically significant difference from zero) decrease in the RR of bacteriologically-confirmed TB prevalence from the first to the third quartile of country- and year-matched ART coverage in our dataset. This is equivalent to a RR of 4·60 (2·42–7·39) at 12% ART coverage (first quartile), and 3·30 (1·79–5·25) at 55% ART coverage (third quartile).

We also estimated differences in the TB notification rate ratio using data matched to the nationally-representative surveys included in primary analysis; this ratio was consistently lower than the TB prevalence risk ratio in the same country (Table 3). Pooled across the five countries included in the notification analyses, the TB notification rate among people living with HIV was over twice those estimated among people without HIV (RR = 2·47, 1·34–3·34).

The estimated ratios of annual TB notifications to TB prevalence (N:P ratios) were higher among people living with vs. without HIV in all examined countries, with pooled N:P ratios of 1·74 (0·59–4·56) and 0·48 (0·17–1·20), respectively. On average, estimated N:P ratios were four times higher in people living with versus without HIV (3·91; 1·36–9·08). Covariate analysis of the differences in N:P ratios at the different ART coverage values produced uncertain estimates. The estimated N:P ratio among people living with HIV was 1·27 (0·51–2·93) at 12% ART coverage and 1·35 (0·56–3·06) at 55% coverage, representing a 7·2% (−56.6%–154·6%) decrease.

Of the 12 surveys included in the primary analysis, four were evaluated to have low, seven were evaluated to have moderate, and one was evaluated to have high risk of bias. Figure S2 shows the distribution of studies across all 8 evaluated criteria.

## Discussion

We analyzed data from 17 TB prevalence surveys conducted in 10 LMICs, to understand systematic differences in TB prevalence in populations with and without HIV. Our primary analysis, using data from 12 surveys in 9 LMICs, found a four-times higher community TB prevalence among people living with HIV compared to people without HIV. We further found that ratios of TB notifications to bacteriologically-confirmed TB prevalence were substantially higher for people living with versus without HIV, suggesting that rates of TB diagnosis are systematically higher among people with HIV. These estimates highlight the elevated burden of undiagnosed TB among people living with HIV.

The finding of elevated TB prevalence among people living with HIV is consistent with previous analyses. Several studies have measured TB prevalence among HIV-positive cohorts, such as persons undergoing screening prior to ART or TB preventative therapy, and reported elevated TB prevalence compared to the general population.^55^ These studies, however, often omit a non-HIV comparator population, requiring comparison to external TB case notifications or prevalence estimates. By using data collected from a community sample, we constructed prevalence risk ratios that limit study-level differences which might confound this relationship. It is important to note that we examined marginal differences in TB prevalence by participant HIV status. In reality, the difference in TB prevalence between people living with and without HIV will be influenced by several demographic and sociocultural factors, such as sex, age, living conditions, socioeconomic status, and healthcare access.^56,57^ Effective intervention design should consider intersections between these factors as well.

Estimated N:P ratios suggested that people living with HIV have a nearly four-fold shorter duration of undiagnosed disease, with an average time to diagnosis of less than one year (signified by an N:P ratio exceeding one). While previous studies in Africa have reported diagnostic delays of 3-8 weeks, these studies typically only include individuals who ultimately receive a diagnosis through patient-initiated care-seeking.^58,59^ This approach is subject to an individual developing symptoms, recall bias, and the omission of those for whom diagnosis is made at death, resulting in mismeasurement of the duration of undiagnosed TB. Our pooled N:P ratios suggest longer durations of undiagnosed disease compared to these studies, both for people living with and without HIV, and are more consistent with modelled evidence accounting for unobserved delays.^60^ Prevalence estimates stratified by both participant symptoms and HIV status were not available for our analysis but could further elucidate the impact of asymptomatic TB on the duration of undiagnosed TB.

We further assessed how ART coverage may affect TB prevalence, estimating a lower (albeit uncertain), excess TB prevalence among people living with HIV with higher levels of ART coverage. This result is consistent with other evidence on how ART affects TB-HIV natural history. ART can reverse HIV-associated immunosuppression and moderate the increased risk of TB among people living with HIV. Additionally, higher ART coverage could reduce TB prevalence by shortening the duration of untreated TB among populations living with HIV, given the frequent clinical appointments and elevated TB awareness within ART programs.^6,8,9^

While TB prevalence surveys provide invaluable data on the distribution of TB across populations, they also provide a healthcare contact for people that might otherwise be unreached by health services. As such, these surveys represent an opportunity to identify and provide education on TB co-morbidities, such as HIV. The coordination and implementation of these surveys, however, requires a large financial and personnel investment, which has limited the widespread adoption of HIV testing as a component of TB prevalence surveys. In our analysis, all identified surveys that provided systematic HIV testing to TB screened participants were located in the WHO Africa region, likely reflecting the disproportionate burden of TB-HIV coinfection in this region.^1^ Of the WHO-designated list of 30 countries with a high dual burden of TB and HIV, 22 are located in the WHO Africa region.^61^ The remaining countries (located in South East Asia and South America) were not represented in our dataset, creating a knowledge gap in the interaction of TB and HIV epidemiology outside of Southern and Eastern Africa. While it is possible that the broad patterns estimated in our analysis also apply in other regions, more evidence is need on the intersection of these two epidemics in settings with concentrated HIV epidemics.

This analysis has several limitations. The surveys identified in this review, particularly the sub-national surveys, varied in their approaches for ascertaining TB and HIV status. In particular, some surveys only assessed HIV status via self-report. We addressed this potential bias by analyzing tested and self-reported HIV cohorts separately. Other potential biases are introduced by approaches used to detect TB, with both TB symptom screening and confirmatory diagnostics shown to have lower sensitivity among people living with HIV.^62^ Only one survey included in our primary analysis required symptoms for TB testing, which limits this potential bias; however, the lower sensitivity of TB diagnostic tests among people living with HIV means that the prevalence ratios estimated in our analysis could underestimate the prevalence ratios for all pulmonary TB disease (including culture-negative TB). Furthermore, given diagnostic difficulties, our analysis excluded surveys of extra-pulmonary TB prevalence, which occur at higher rates in people living with HIV.^63^ As such, we likely underestimate the difference in total TB prevalence between people living with and without HIV.^3^ As screening and testing for extra-pulmonary TB in community settings improves, these estimates should be revisited. Finally, as WHO does not report TB notifications among persons with HIV stratified by age group or diagnostic criteria (i.e. bacteriologically-confirmed, smear-positive, etc.), our notification-to-prevalence (N:P) ratios used all TB notifications reported for each country-year pair. As such, our N:P ratios may overestimate the diagnostic probabilities in both persons with and without HIV.

Despite the widely-accepted relationship between HIV and TB, we identified few TB prevalence surveys reporting HIV-disaggregated estimates. This evidence is concentrated in countries with generalized HIV epidemics in Southern and Eastern Africa, yet countries with high TB-HIV burden are distributed globally. We estimated four-times higher risk of bacteriologically-confirmed TB prevalence among people living with versus without HIV. Our findings highlight the elevated risk of TB among people living with HIV as well as the current geographic-imbalance in TB-HIV investigations, emphasizing the need for continued investment in the prevention and detection of TB-HIV co-infection.

## Contributors statement

NAM, PM, and KCH conceptualized the study. NAS, PM, KCH, and NAM developed the study methodology. NS, NAS, AM, HC, MHC, DKR, KCH, PM, and NAM compiled study data. NAS implemented data cleaning and meta-analysis. NAS, PM, KCH, and NAM interpreted results. NAS drafted the manuscript. All coauthors edited, reviewed, and approved the final submitted manuscript. NAS and PM accessed and verified the data. All authors had full access to all the data in the study and accept responsibility for the decision to submit for publication.

## Acknowledgements

We thank Stephanie Su for assistance in study coordination and Dr. Jeff Imai-Eaton for his assistance in collating the UNAIDS data used in the analysis. PM was funded by Wellcome (304666/Z/23/Z) and an NIHR Global Health Research Professorship (NIHR304311). The views expressed are those of the author(s) and not necessarily those of the NIHR or the Department of Health and Social Care. For the purpose of open access, the authors have applied a CC BY public copyright license to any Author Accepted Manuscript version arising from this submission. KCH is supported by the UK FCDO (Leaving no-one behind: transforming gendered pathways to health for TB) and the U.S. National Institutes of Health (R-202309-71190). This research has been partially funded by UK aid from the UK government (to KCH); however the views expressed do not necessarily reflect the UK government’s official policies. The funders of the study had no role in study design, data collection, data analysis, data interpretation, or writing of the report.

## Data sharing statements

All data and code used in this analysis can be found at https://github.com/nswartwo/tb-lmic-prevalence-review and archived on Zenodo (DOI to be added upon acceptance).

